# Sex-Based Utilization of Guideline Recommended Statin Therapy and Cardiovascular Disease Outcomes: A Primary Prevention Healthcare Network Registry

**DOI:** 10.1101/2024.01.22.24301511

**Authors:** Agnes Koczo, Adipong Brickshawana, Jianhui Zhu, Floyd Thoma, Malamo Countouris, Kathryn Berlacher, Martha Gulati, Erin D Michos, Steven Reis, Suresh Mulukutla, Anum Saeed

**Author notes:** Address for Correspondence: Anum Saeed, MD, 3550 Terrace St, Suite 401 Scaife Hall, Pittsburgh, PA 15216 E, Twitter: @AnumSaeedMD. Co-first authors (Agnes Koczo, Adipong Brickshawana).

## Abstract

**Background:** In the US, women have similar cardiovascular death rates than men. Less is known about sex differences in statin use for primary prevention and associated atherosclerotic cardiovascular disease (ASCVD) outcomes.

**Methods:** Statin prescriptions using electronic health records were examined in patients without ASCVD (myocardial infarction (MI), revascularization or ischemic stroke) between 2013-2019. Guideline-directed statin intensity (GDSI) at index and follow-up visits were compared among sexes across ASCVD risk groups, defined by pooled-cohort equation. Cox regression hazard ratios (HR) [95% CI] were calculated for statin use and outcomes (myocardial infarction, stroke/transient ischemic attack (TIA), and all-cause mortality) stratified by sex. Interaction terms (statin and sex) were applied.

**Results:** Among 282,298 patients, (mean age ∼ 50 years) 17.1% women and 19.5% men were prescribed any statin at index visit. Time to GDSI was similar between sexes, but the proportion of high-risk women on GDSI at follow-ups was lower compared to high-risk men (2-years: 27.7 vs 32.0%, and 5-years: 47.2 vs 55.2%, p<0.05). When compared to GDSI, no statin use was associated with higher risk of MI and ischemic stroke/TIA amongst both sexes. High-risk women on GDSI had a lower risk of mortality (HR=1.39 [1.22-1.59]) versus men (HR=1.67 [1.50-1.86]) of similar risk (p value interaction=0.004).

**Conclusion:** In a large contemporary healthcare system, there was underutilization of statins across both sexes in primary prevention. High-risk women were less likely to be initiated on GDSI compared with high-risk men. GDSI significantly improved the survival in both sexes regardless of ASCVD risk group. Future strategies to ensure continued use of GDSI, specifically among women, should be explored.

## Introduction

Atherosclerotic cardiovascular disease (ASCVD) is the number one cause of cardiovascular death in both women and men in the United States.^1^ Since the 1990s, there has been a steady decline in the number of cardiovascular disease deaths.^1^ This may be in part due to improved care of patients with acute coronary syndrome, but also due to greater effort at primary and secondary prevention of ASCVD.^2^ An integral part of ASCVD prevention is the use of statin therapy to treat hyperlipidemia. Statins are a cornerstone therapy effective at reducing ASCVD risk and events. ^2–5^ Nevertheless, underutilization of statin therapy remains prevalent in everyday practice leading to missed opportunities to reduce ASCVD and mortality. ^6^

The decline in cardiovascular disease death over the past several decades has not been equally shared between women and men. Since 1984, ASCVD mortality had been higher in women as compared to men. ^7^ More recently, total cardiovascular deaths have been similar in women as in men (for men n=487,188 and for women n=441,525 in the year 2020), with recent upward trends in cardiovascular death for both men and women. ^8^

As detailed by two recent American Heart Association (AHA) Scientific Statements, there are distinct differences in the pathophysiology, presentation, and outcome of cardiovascular disease between women and men.^7,9^ Particularly, in the setting of ischemic heart disease, women are more likely to present with additional non-chest pain symptoms and at an older age compared to men. The risk of ASCVD does increase comparably between men after age of 45 years and women after age of 55 years.^77^There is an increasing body of literature noting sex-specific risk factors, particularly among women, including condition in and around pregnancy and menopause. Pregnancy associated cardiovascular risk factors include history of infertility or use of reproductive assistive technology, hypertensive disorders of pregnancy, spontaneous preterm birth, both small and large for gestational age babies, low birth weight, placental abruption and gestational diabetes.^10^ Menopause associated cardiovascular risk factors include early or premature surgical or natural menopause as well as extended or late postmenopausal hormone therapy use.^11,12^ Non-traditional cardiac risk factors including polycystic ovary disease as well as autoimmune disease associated with increased cardiovascular event (rheumatoid arthritis and lupus) are also disproportionately or elusively impacting women.^13,14^ Some of these risk factors have already been incorporated in the American College of Cardiology/American Heart Association (ACC/AHA) 2018 Cholesterol Management Guideline as ASCVD risk enhancers.^3^

In spite of sex-specific risk enhancers, the use of statins have been shown to be equally effective and safe in women and men, resulting in similar reductions in coronary events, coronary revascularization, and stroke. ^15^Nevertheless, several prior studies have shown that women with established ASCVD are less likely to be on statins, stay adherent to statins, and to achieve intended low-density lipoprotein (LDL) cholesterol goals while on statins.^16–19^ This likely contributes to the difference in decline of cardiovascular disease between sexes since initiation of statin use. Notably, women with diabetes received less intense dyslipidemia treatment compared to men, and women with established cardiovascular disease were less likely to have adequate LDL control.^16,20^ Most of these studies were done in patients with established ASCVD and for secondary prevention. Data regarding sex-differences across longitudinal follow-up with statin use in primary prevention, specifically in a real-world community-based setting, and its relationship to outcomes has not prior been reported.

In this study, we assessed whether sex-based differences exist in statin utilization for primary prevention across the ACC/AHA risk-based categories in a large health care network at the University of Pittsburgh Medical Center (UPMC) across longitudinal follow-up. Using electronic health record (EHR) data, we assessed the resulting effects of statin use and guideline-directed statin intensity (GDSI) and the association with ASVCD outcomes (myocardial infarction (MI), stroke, and mortality) comparing men and women.

## Methods

A detailed description of the methods used in creation and definitions of the primary prevention cohort across our healthcare system has been published previously.^6^ Briefly, the primary prevention cohort consisted of men and women ages 20 to 79 years. Participants were evaluated in at least two health care interactions between January 2013 and December 2017 in the University of Pittsburgh Medical Center (UPMC) health care system. UPMC is a large multihospital network comprised of 400 outpatient clinics and nearly 40 hospitals based in Pittsburgh, PA and spanning across Pennsylvania. Patient were censored following December 2017 for inclusion in study and event were collected up to December 2020 for a median of 6 years of follow-up. All included patients had at least one lipid profile drawn within 180 days of the first interaction. Data was collected via review of the patients’ EHR records. Exclusion criteria included prior coronary artery disease (ICD-9: angina, myocardial infarction, revascularization, and ischemic cardiomyopathy), cerebrovascular disease or stroke (ICD-9 and ICD-10: transient ischemic stroke, ischemic stroke, and peripheral artery disease), history of rhabdomyolysis, and being in a skilled nursing facility or hospice. A total of 2,348,822 patients were evaluated in the UPMC health care system during the study period, and 282,298 patients (12%) met these inclusion criteria for the study.

Eligible patients in this primary prevention cohort ^6^ were all divided at their index visit based on their sex (in this manuscript: female denoted as women and male as men, EMR has limited data on self-identified gender during these time periods), pooled-cohort equations estimated 10-year ASCVD risk (low: <5%, borderline: 5%-7.4%, intermediate: 7.5%-19.9%, and high: ≥20%), ^21^ and statin prescription. Guideline-directed statin intensity (GDSI) was as defined by the 2013 and 2018 ACC/AHA cholesterol guidelines.^3,22^ Less than GDSI (<GDSI) was defined as statin use of lower intensity than indicated by the guidelines. For example, GDSI was defined as “at least moderate” intensity for intermediate and high intensity statin for high-risk. Statin intensity was further defined by statin type and dosing. This included low (simvastatin 10 mg, lovastatin 20mg, fluvastatin 20 and 40 mg), moderate (atorvastatin 10 and 20 mg, rosuvastatin 5 and 10 mg, simvastatin 20 and 40 mg, lovastatin 40 and 80 mg), or high (atorvastatin 40 and 80 mg, rosuvastatin 20 and 40 mg) statin intensities. ^23^ Of note, patients on pravastatin represented less than 0.001% of the cohort, so they were excluded from the study. Time to GDSI was defined as years from first health care encounter to achieving GDSI.

Incident ASCVD events that were assessed included ischemic stroke/transient ischemic attack (TIA), myocardial infarction (MI), and mortality. All outcomes were surveyed and included as defined per the ICD-9 and ICD-10 coding in the EHR. Mortality was assessed using the United States Social Security Death Index. Our health care system is exempt from the 3-year delay period by the Social Security Administration.

### Statistical Analysis

Baseline descriptive statistics of the study sample for each continuous variable and frequency tables for each categorical variable were initially analyzed to detect outliers and missing values. Missing data were uncommon and, where applicable, were replaced by the simple mean imputation across the risk groups. Descriptive characteristics were normally distributed and were presented as mean and standard deviation (SD) for continuous variables, and frequencies and proportions for categorical variables. The difference of means across the ASCVD groups was assessed by 1-way ANOVA, while the difference of frequencies was compared using the χ2 test. All analyses were completed using SAS version 9.4 software (SAS Institute). Statistical significance was set at αLJ=0.05. All tests of statistical significance were 2-tailed. SAS was used to calculate mean time to GDSI amongst each group. The primary and composite outcome incident rates (IRs) and mortality were calculated as event rates per 1000 person-years across risk categories stratified by statin utilization. The 95% CIs were estimated using a generalized linear model with the Poisson distribution. Further, Cox proportional hazards regression models were used to compare the hazard ratios of primary outcomes among each risk category and between use of GDSI versus and no statin use before the first event. The survival curves for cardiovascular outcomes and mortality comparing the statin therapy groups were plotted after Cox proportional hazards regression adjusted for mean values of age, high-density lipoprotein cholesterol (HDL-C), low-density lipoprotein cholesterol (LDL-C), systolic blood pressure at index visit, and reference groups of categorical variables (white self-identified race, men, current smoker and no diabetes). The GDSI variable was treated as time invariant, and classification of GDSI was based on the statin status before each outcome. Individuals without the primary outcome (composite ASCVD events and mortality) were censored between 2 and 7 years after baseline in the corresponding analysis. The time to GDSI across ASCVD risk groups was estimated using the Kaplan-Meier method.

## Results

### Baseline Demographics

The baseline characteristics for women (n= 159,100) and men (n= 123,198) (mean age ∼ 50 years) are shown in Table 1. In general, fewer women were noted to have diabetes (9.6 vs. 11.0%), hypertension (22.5 vs 27.8%), and active smoking status (22.4 vs 26.0%) as compared with men, equating to a lower Elixhauser (ELIX) comorbidity score (ELIX reference). However, women were marginally older (50.6 vs 49.5 years) and more likely to self-identify as Black (7.7 vs 6.7%). The index visit LDL-c (114 ± 32.4 mg/dL in women, 115 ± 31.5 mg/dL in men) and total cholesterol (196 ± 36.9 mg/dL in women and 191 ± 36.1 mg/dL in men) levels were comparable between sexes. A lower percentage of women were prescribed any statin compared with men (17.1 vs. 19.5%) overall. (**Table 1**)

**Table 1.**
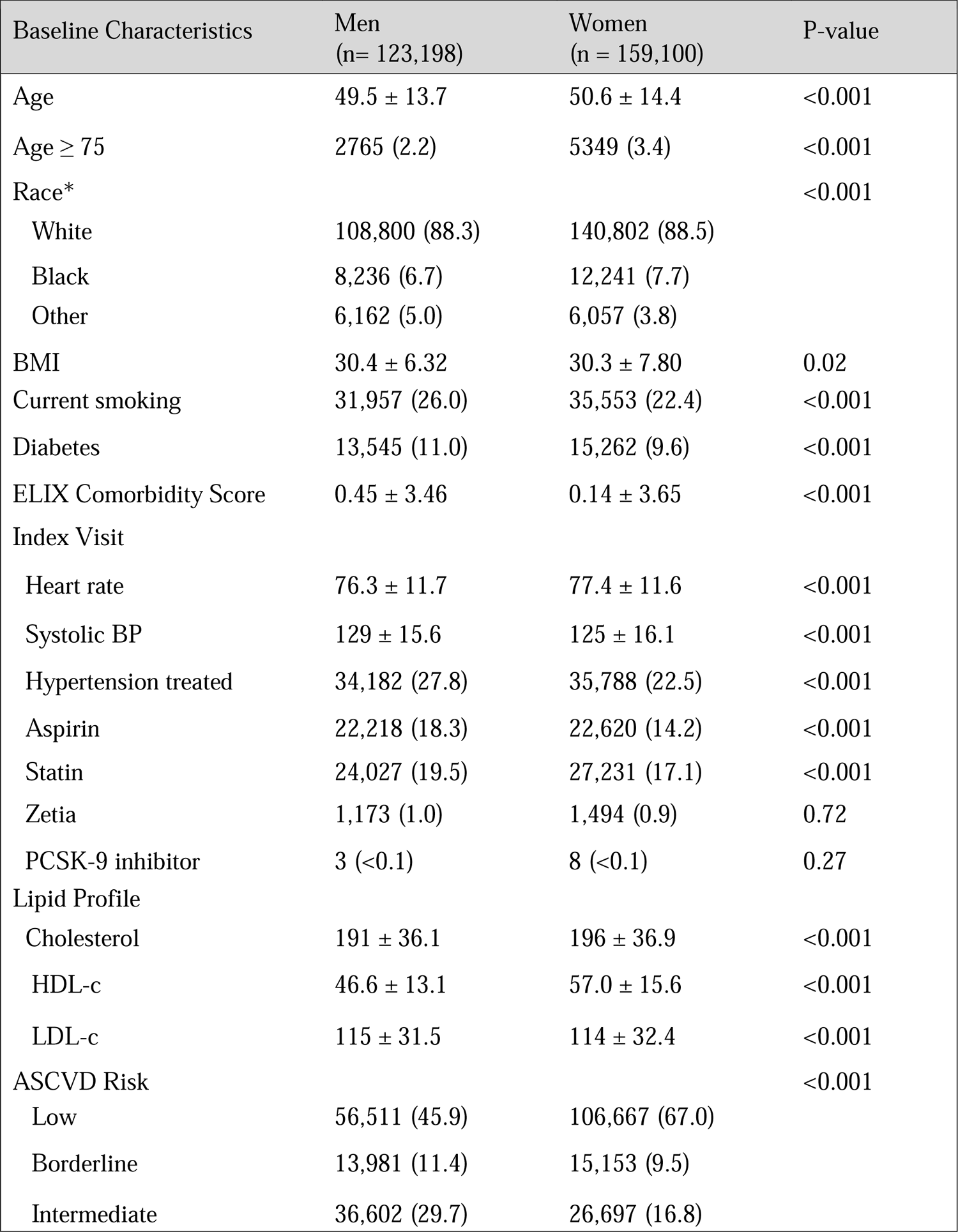
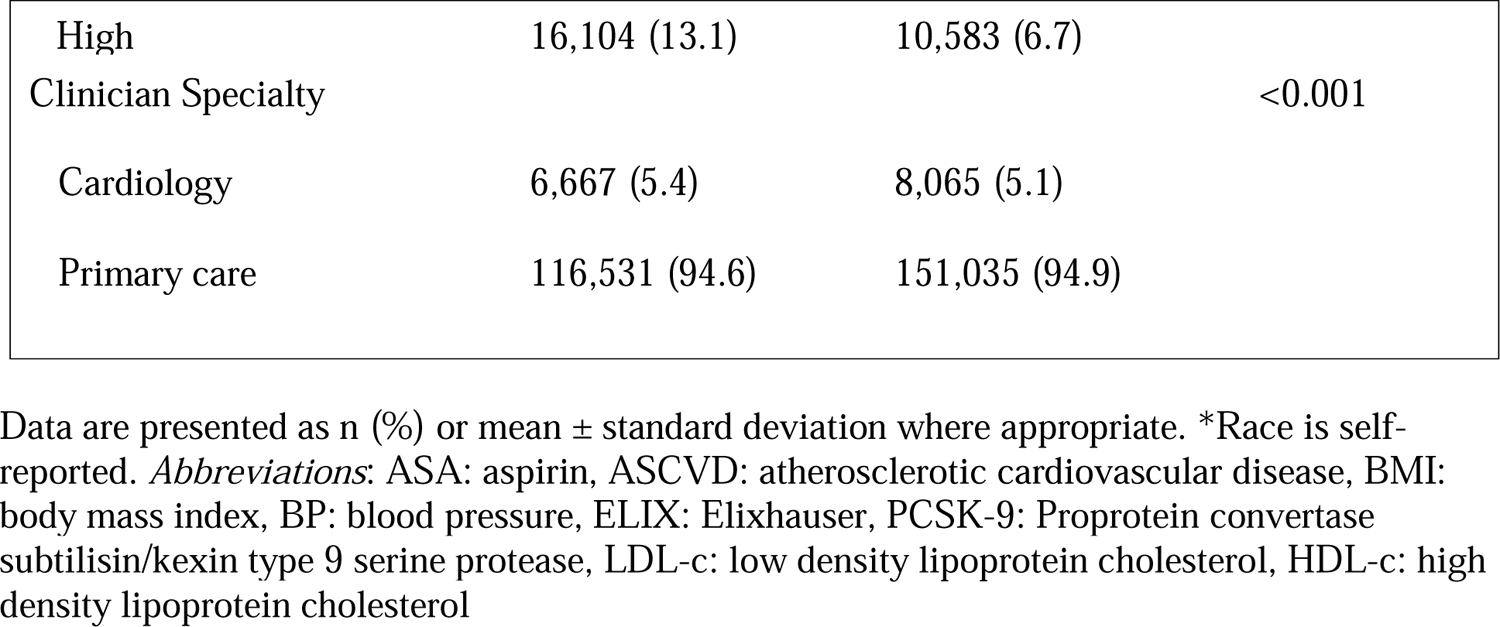
Sex Differences in Baseline Demographics.

### LDL-C by Sex and ASCVD risk

At baseline, LDL-C levels among both intermediate ASCVD risk men and women were higher in those not on statin (men: 124.2, women: 128.9mg/dL) as compared to those on GDSI (men: 105.3, women 106.3 mg/dL). This was consistent among high ASCVD risk men and women (men no statin: 119.7, women no statin 126.1 mg/dL; men GDSI: 98.9, women GDSI: 99.7 mg/dL). (**Figure 1**) At the censor/final data collection timepoint, LDL-C levels among both intermediate and high ASCVD risk men and women were similarly higher in those not on statin (intermediate no statin men: 118.7, intermediate no statin women: 122.2 mg/dL; high risk no statin men 113.6, high risk no statin women 120.2) as compared to those on GDSI (intermediate GDSI men: 119.7, intermediate GDSI women 120.7 mg/dL; high risk GDSI men 110.3, high risk GDSI women 111.5 mg/dL). (**Supplemental Table I**)

**Figure 1.**
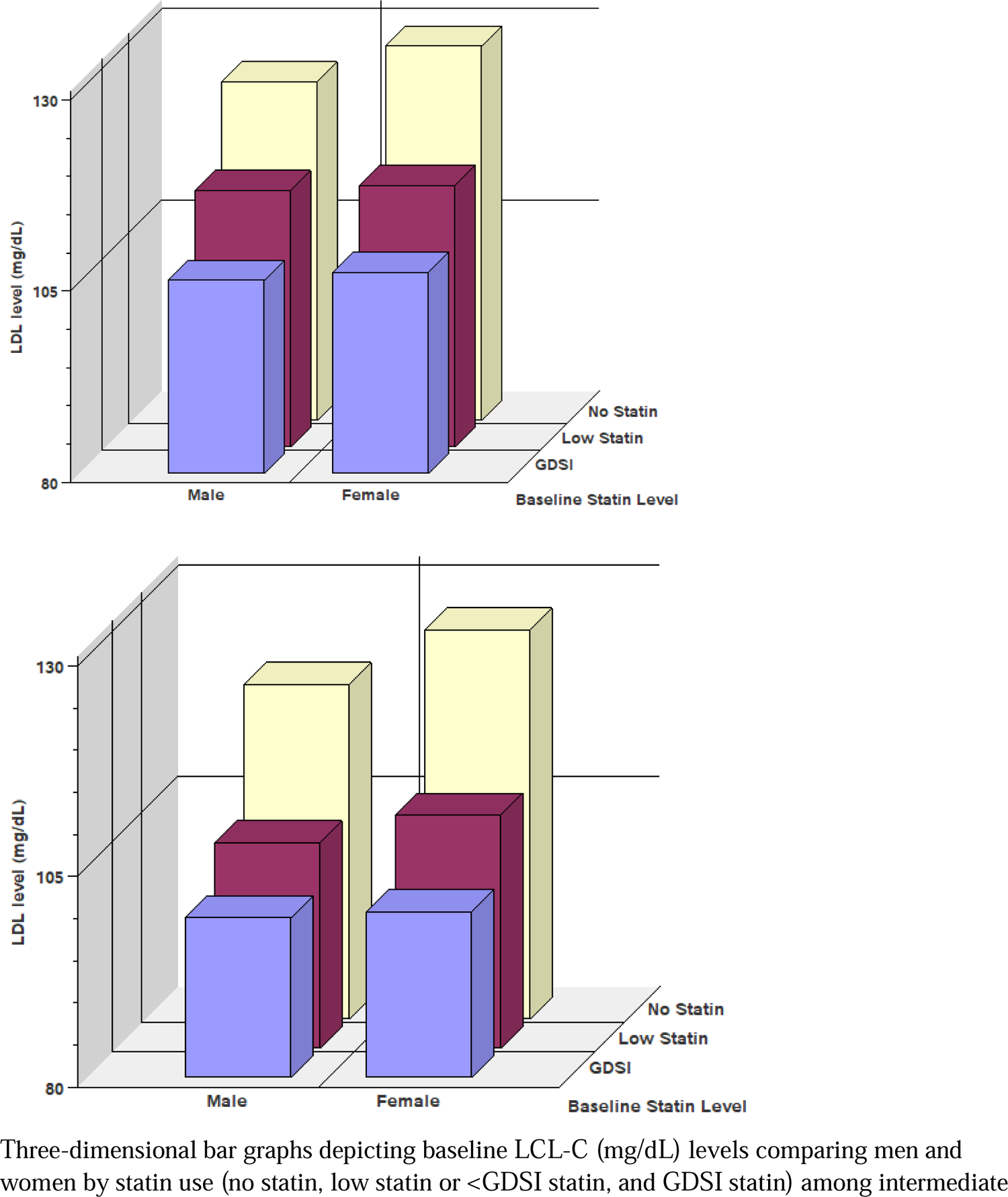
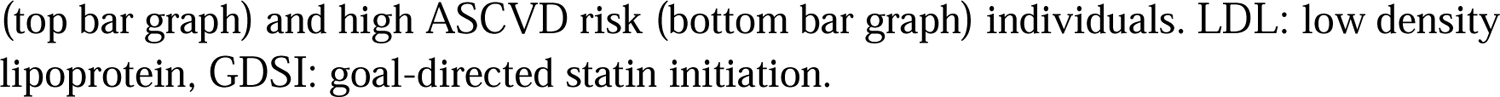
Baseline LDL-C levels by Sex and Statin Use Among Intermediate and High ASCVD Individuals.

### Statin Use by Sex and ASCVD Risk

When stratified by the ASCVD 10-year risk score, there were more women in the low-risk group (67.0 vs 45.9%) compared with men (**Table 1**). Conversely, there were more men in the borderline (11.4 vs 9.5%), intermediate (29.7 vs 16.8%), and high (13.1 vs 6.7%) risk groups compared with women.

Although there were less women in the high-risk group, the percentage on any statin as well as GDSI was greater at index visit as compared to men (high risk. any statin: 45.6 vs 41.6%, high risk, GDSI: 41.7 vs 38.8%). More than half of high-risk patients among both sexes were not prescribed any statin at index visit (men: 58.4 vs. women: 54.4%).At follow-up visits, the percentage of high-risk women on GDSI was significantly lower compared with men (High Risk GDSI: 63.8 vs. 66.9%). (Table 2)

**Table 2.** Proportion of Statin Therapy Utilization On Index and Follow-up Visits Across Intermediate and High ASCVD Risk Categories.

| Index Visit |  |  |  |  | Follow-up |  |  |  |
| --- | --- | --- | --- | --- | --- | --- | --- | --- |
| ASCVD Risk | Intermediate |  | High |  | Intermediate |  | High |  |
| Sex (n) | Men<br>(36,602) | Women<br>(26,997) | Men<br>(16,104) | Women<br>(10,583) | Men<br>(36,602) | Women<br>(26,997) | Men<br>(16,104) | Women<br>(10,583) |
| No statin | 26,661<br>(72.9) | 17,597<br>(65.9) | 9,403<br>(58.4) | 5,760<br>(54.4) | 16,627<br>(45.4) | 10,731<br>(40.2) | 4,882<br>(30.3) | 3,418<br>(32.3) |
| < GDSI | 730<br>(2.0) | 730<br>(2.7) | 449<br>(2.8) | 408<br>(3.9) | 773<br>(2.1) | 890<br>(3.3) | 457<br>(2.8) | 411<br>(3.9) |
| GDSI | 9,621<br>(25.3) | 8,370<br>(31.4) | 6,252<br>(38.8) | 4,415<br>(41.7) | 19,202<br>(54.5) | 15,076<br>(56.5) | 10,765<br>(66.9) | 6,754<br>(63.8) |

Data presented as n (%) of patients on targeted treatment based on risk group at index visit and the highest treatment they ever received during follow-up period. \*P < 0.05. Abbreviations: ASCVD- atherosclerotic cardiovascular disease, GDSI- guideline-directed statin intensity.

### Statin Use by Sex and Time to GDSI

Evaluating the patients over time, the mean times to GDSI for women and men in both the intermediate and high-risk groups were not different (**Table 3**). However, based on the Kaplan-Meier probability estimates, high-risk women were persistently less likely to be on GDSI compared to high risk-men both at 2- and 5-years follow-ups (2-years: 27.7 vs 32.0%, and 5-years: 47.2 vs 55.2%, p<0.05) **(Supplemental Table II)** (**Figure 2**).

**Figure 2.**
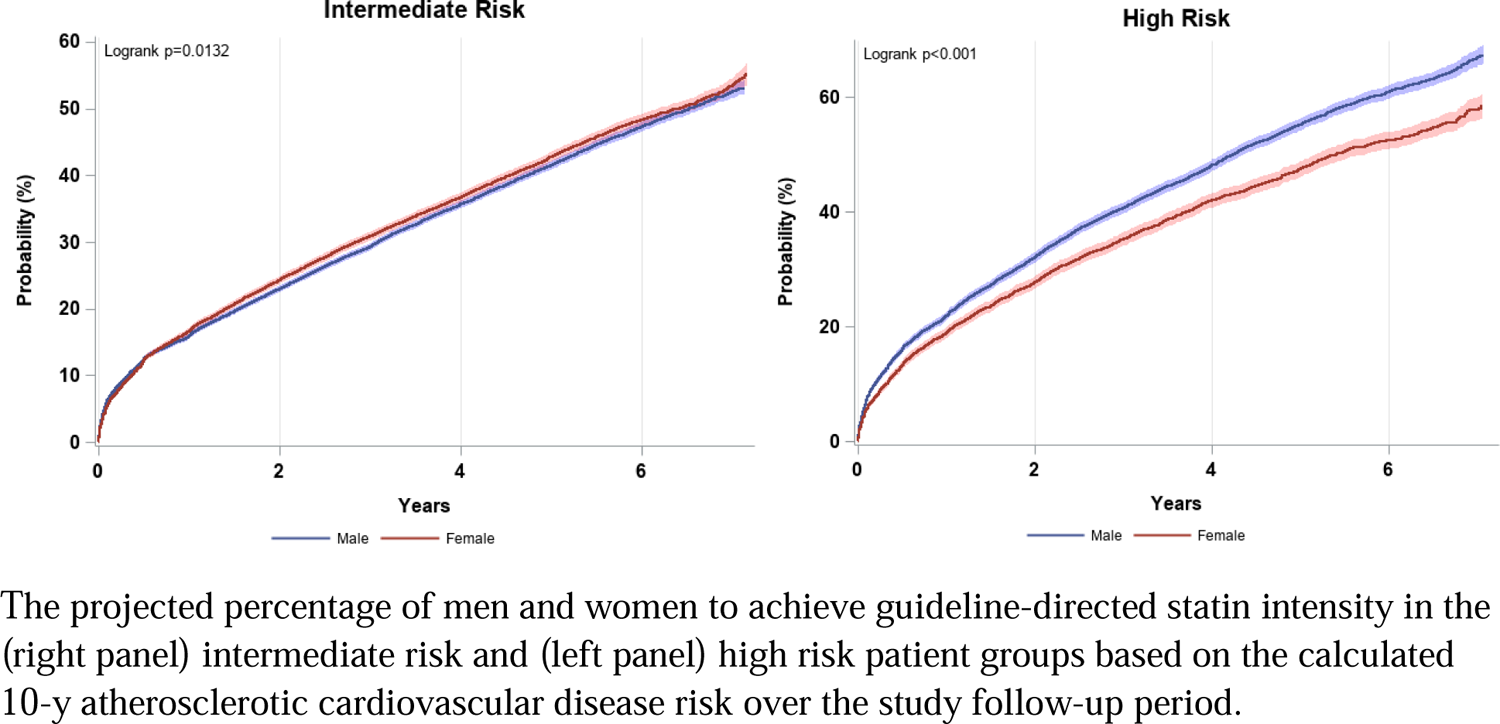
Probability of GDSI Use Over Follow-Up Period

**Table 3.** Time to Guideline Directed Statin Intensity Initiation During Follow-up.

| ASCVD Risk | Statin | Time to GDSI (months) |  | P value |
| --- | --- | --- | --- | --- |
|  |  | Men | Women |  |
| Intermediate | Moderate | 20.7±20.8 | 20.8±20.4 | 0.04* |
|  | High | 27.3±21.7 | 26.8±21.6 | 0.3 |
| High | Moderate | 19.1±19.6 | 19.5±19.6 | 0.3 |
|  | High | 24.8±21.0 | 24.8±21.7 | 0.7 |

Data are presented as mean ± SD time (in months) to initiation of intermediate or high intensity statin therapy for men and women without statin prescription at index visit.
. \*P = 0.05. ASCVD: atherosclerotic cardiovascular disease, GDSI: guideline-directed statin intensity. The ASCVD risk categories are defined per the pooled-cohort equations based on 10-year ASCVD risk calculator. \*P <0.05.

### Hazard Ratio of Event by Sex and Statin Use

Over a median follow-up of 6 years, no statin use was associated with a significantly higher risk of adverse cardiovascular events as compared with GDSI (MI: HR_Women_ = 1.32 [1.07-1.62] vs HR_Men_ = 1.46 [1.25-1.72]), stroke/TIA: (HR_Women_ = 1.61 [1.31-1.98] vs HR_Men_ = 1.63 [1.36-1.96]) amongst both high-risk men and women. GDSI use (compared to no GDSI) in high-risk women had lower incident all-cause mortality (HR_Women_ = 1.39 [1.22-1.59]) compared with men counterparts (HR_Men_ = 1.67 [1.50-1.86]) *(p-value interaction = 0.004*). (**Table 4**). Similarly, in the intermediate risk group, patients of both sexes on no statin also had a higher risk of MI, stroke, and mortality compared to those on GDSI. Overall, GDSI significantly improved the survival in both sexes regardless of ASCVD risk group.

**Table 4.** Sex Differences in Adverse Outcomes Comparing on No Statin Among Intermediate and High-Risk Individuals.

| Events | Men |  | Women |  |
| --- | --- | --- | --- | --- |
|  | Intermediate risk | High risk | Intermediate risk | High risk |
| MI | 1.59<br>(1.38 – 1.84) *** | 1.46<br>(1.25 – 1.72) *** | 1.54<br>(1.30-1.84) *** | 1.32<br>(1.07-1.62)** |
| Stroke-TIA | 1.32<br>(1.09-1.60) ** | 1.63<br>(1.36-1.96) *** | 1.72<br>(1.42-2.08) *** | 1.61<br>(1.31-1.98) *** |
| Mortality | 1.67<br>(1.50-1.86) *** | 1.67<br>(1.50-1.86) *** | 1.65<br>(1.47-1.85) *** | 1.39<br>(1.22-1.59) *** |

Hazard Ratios using guideline-directed statin intensity as reference. Data presented as median (IQR). Values expressed as hazard ratio (95% CI). \*P<0.05, \*\* P<0.01, \*\*\*P<0.001. TIA: transient ischemia attack, MI: myocardial ischemia

## Discussion

In this study of sex-based assessment of statin utilization and ASCVD outcomes in a contemporary primary prevention cohort, stratified by 10-year ACC/AHA risk, we present three key findings; 1) high risk women were persistently less likely to be on GDSI compared with high risk men both at 2 and 5 years follow-up (2-years: 27.7 vs 32.0%, and 5-years: 47.2 vs 55.2%) despite comparable risk and higher proportion of GDSI at baseline, 2) both intermediate and high risk women and men had lower risk of MIs, stroke and mortality when on GDSI compared to when on no statin and 3) GDSI significantly improved the survival in both sexes across all ASCVD risk groups.

Several prior studies have shown disparities in statin utilization among women compared with men for secondary prevention of ASCVD events. ^16–20,24^ Our study is novel as 1.) we demonstrate sex-based differences in statin use and ASCVD outcomes in a purely primary prevention cohort from a real world, large contemporary healthcare systems in the United States and 2.) our study examined longitudinal outcomes including Social Security Index verified all-cause mortality. Further, our data were strengthened by statin associated ASCVD outcomes as well as social security death index adjudicated mortality.

In limited studies performed in the United States evaluating primary prevention patients among registry enrolled participants without longitudinal outcomes, differences comparing men and women regarding statin use are less clear with mixed results. ^25,26^ Navar et al. have previously reported that women were less likely than men to receive guideline-recommended statin intensity therapy in the PALM registry.^27^ Although the PALM registry is also a contemporary sample, the number and percentage of patients in primary prevention were far less than this study. Further, our study represents a younger average patient population. Further, recall bias in the survey-based registry as well as a shorter duration of enrollment (all within 2015 in PALM versus 2013-2017 in the current study) may be some of the reasons for differing results.^27^

In regards to longitudinal statin use, we found that the probability of high-risk women being on GDSI was lower as compared to high-risk men persistently at 2- and 5-year follow-up. Prior data focused on evaluating reasons for sex differences in guideline-directed statin therapy initiation.^27^ Our data adds to our understanding of sex differences over time following the introduction of the 10-year risk estimation in 2013. Despite sex-specific risk enhancers added to the guidelines, high-risk women had comparatively lower initiation of guideline-concordant statin intensity over time as compared to high-risk men. Such is illustrated by a larger proportion of high-risk women on less than guideline directed statin therapy compared to men during the follow up period. It is also important to note that the ASCVD risk calculation may not fully capture sex-specific risk factors, some of which are now emphasized as risk-enhancers in the most recent guidelines.^\^ ^3^ Given more sex-specific non-traditional risk factors are attributed to women, there may be an overall underestimation of women’s risk when looking at ASCVD calculations in isolation.

Although we did not evaluate the reasons for differences in sex associated differences in in our data, prior studies have shown that both clinician-driven and patient-driven factors may contribute to this disparity in statin utilization.^27^ Clinician-driven factors may include clinicians’ sex-specific biases as well as an underappreciation of women’s true ASCVD risk. In the PROMISE trial, diagnostic workup for possible coronary artery disease (CAD) was shown to be biased by the patient’s sex. ^28^ Another study showed clinicians were more likely to consider men to be of higher risk compared women with an identical clinical scenario and consequently were more likely to prescribe lipid-lowering medications to men. ^29^ Incidence of risk factors for clinical ASCVD increase in the post-menopausal period, and risk among women to match their men counterparts in later life. A lack of knowledge about non-traditional cardiovascular risk factors including adverse pregnancy outcomes as well as early menopause may lead to under prescribing of GDSI. From the patient’s perspective, women of child-bearing status may have concerns about statins and teratogenicity. It’s important to note the FDA has removed the category X classification for statins during pregnancy.^30^ Further, certain statins have been tested to treat hypertensive disorders during pregnancy.^31^ In addition, women have tended to report more adverse effects ascribed to statin.^24,32^ Prior studies have hypothesized this could be due to differential exposure to public warnings affecting patients’ perceptions as well as possible drug-drug interactions.^24^

While sex differences in statin utilization were an important focus on this study, it’s important to underscore missed opportunities for statins among both sexes. In particularly, over 50% of high-risk patients among both sexes were not prescribed any statin at index visit. Further, the average times to reach appropriate intensity statin was 20-27 months for both sexes. These data highlight tremendous opportunities to effectively risk stratify and initiate statin among providers for both sexes.

This study had several limitations. First, all EMR data related limitations apply including but not limited to data entry inaccuracy, over/inaccurate coding, misclassifications of medication exposure as well as non-adjudicated endpoints. As reported previously, these should be applied to our study. ^33^ However, all-cause mortality was reported using an SSI validated measure in our outcomes. Furthermore, racial and ethnic diversity in our patient population was limited. Self-identified gender was not effectively documented in the EMR during time period of the study and limits our assessment of gender differences in statin utilization and outcomes. In addition, any treatment for ASCVD or statin prescription at outside UPMC sites could not be properly assessed. Further, EMR data limits detailed analysis on cross-over from different statin intensity groups. Despite this, data from the large, multicenter UPMC system throughout diverse socioeconomic areas of Pennsylvania represents a strength of these findings among other single center or single system studies. Finally, we did not have socioeconomic data or insurance status of patients which may have factored into statin non-adherence.

In spite of differences in statin prescriptions, both men and women in intermediate and high ASCVD risk groups were shown to benefit from a reduction in future atherosclerotic cardiovascular events and mortality when on GDSI compared to no statin. This is consistent with prior literature. ^15,34^ Collectively, these findings underscore the importance for clinicians and patients to be cognizant of both the underutilization of statin in all patients and for continued risk assessment for initiation of statins on follow-up, particularly among women, for optimal prevention of ASCVD events.

## Conclusion

In summary, this study showed that while women received GDSI comparatively more than men at index visit, fewer high risk women were prescribed GDSI for primary prevention on follow-up. Additionally, although the time to achieving GDSI was comparable between sexes, high-risk women were less likely to be on GDSI at 2- and 5-years of follow up. Lastly, we showed that intermediate and high-risk individuals of both sexes had lower risk of MIs, stroke, and death when on GDSI compared to no statin. Future efforts should focus on initiatives to improve guideline implementation for optimal statin utilization to prevent cardiovascular events in both sexes.

## Abbreviations

ASCVD: Atherosclerotic cardiovascular disease

AHA: American Heart Association

EHR: Electronic health record

GDSI: Guideline-directed statin intensity

MI: Myocardial infarction

## Data Availability

Data may be provided upon reasonable requests.

