## Supplemental Table for "Sex-Based Utilization of Guideline Recommended Statin Therapy and Cardiovascular Disease Outcomes: A Primary Prevention Healthcare Network Registry"

**Supplemental Tables**

| ASCVD Risk | Sex | Statin Use Group | Patient (n) | Mean LDL-C (mg/dL) | Std Dev |
| --- | --- | --- | --- | --- | --- |
| Intermediate | Men | GDSI | 9261 | 105.3 | 33.1 |
|  |  | < GDSI | 730 | 113.5 | 30.8 |
|  |  | No statin | 26610 | 124.2 | 32.2 |
|  | Women | GDSI | 8370 | 106.3 | 36.3 |
|  |  | < GDSI | 730 | 114.2 | 30.9 |
|  |  | No statin | 17597 | 128.9 | 33.6 |
| High | Men | GDSI | 6251 | 98.9 | 33.5 |
|  |  | < GDSI | 449 | 104.4 | 32.8 |
|  |  | No statin | 9403 | 119.7 | 33.2 |
|  | Women | GDSI | 4415 | 99.7 | 34.7 |
|  |  | < GDSI | 408 | 107.7 | 30.1 |
|  |  | No statin | 5760 | 126.1 | 35.5 |

| ASCVD Risk | Sex | Statin Use Group | Patient (n) | Mean LDL-C (mg/dL) | Std Dev |
| --- | --- | --- | --- | --- | --- |
| Intermediate | Men | GDSI | 19201 | 119.7 | 36.55 |
|  |  | < GDSI | 773 | 118.0 | 31.91 |
|  |  | No statin | 16627 | 118.7 | 29.49 |
|  | Women | GDSI | 15076 | 120.7 | 39.16 |
|  |  | < GDSI | 890 | 122.8 | 34.24 |
|  |  | No statin | 10731 | 122.2 | 31.07 |
| High | Men | GDSI | 10764 | 110.3 | 37.08 |
|  |  | < GDSI | 457 | 107.6 | 32.93 |
|  |  | No statin | 4882 | 113.6 | 29.17 |
|  | Women | GDSI | 6754 | 111.5 | 39.31 |
|  |  | < GDSI | 411 | 113.6 | 33.66 |
|  |  | No statin | 3418 | 120.2 | 32.70 |

**Supplemental Table I**. Low-density lipoprotein concentration (LDL-C) level differences stratified by sex, ASCVD risk, and statin use (no statin, <GDSI statin, and GDSI statin).  LDL-C levels in the full cohort based on statin intensity at 1.) index time point (top table) and 2.) Censor/final date (bottom table) stratified by ASCVD risk. Data is presented as mean ± standard deviation. ASCVD: atherosclerotic cardiovascular disease.

| Estimated Probability of GDSI at Follow-ups | | | |
| --- | --- | --- | --- |
| ASCVD risk | Sex | 2 years | 5 years |
| Intermediate | Men | 23.0 (22.5-23.4) | 41.4 (40.8-42.1) |
|  | Women | 24.2 (23.6-24.8) | 42.8 (42.0-43.5) |
| High | Men | 32.0 (31.1-32.9) | 55.2 (54.1-56.3) |
|  | Women | 27.7 (26.7-28.8) | 47.2 (46.1-48.8) |

**Supplemental Table II. K-M Estimated Probability of Patients on GDSI Therapy At 2- and 5-Year Follow-ups.** Data is presented as median (IQR).

| ASCVD Risk | Total  (159,100) | Low  (106,667) | Borderline  (15,153) | Intermediate  (26,697) | High  (10,583) | P-Value |
| --- | --- | --- | --- | --- | --- | --- |
| Age | 50.6 ± 14.4 | 44.3 ± 11.5 | 57.9 ± 9.53 | 63.3 ± 10.4 | 70.4 ± 10.8 | <0.001 |
| Age ≥ 75 | 3.4 | 0.0 | 0.0 | 3.7 | 41.2 | <0.001 |
| Race |  |  |  |  |  | <0.001 |
| White | 88.5 | 88.4 | 90.6 | 88.5 | 86.9 |  |
| Black | 7.7 | 6.9 | 7.1 | 9.6 | 11.4 |  |
| Other | 3.8 | 4.7 | 2.3 | 2.0 | 1.7 |  |
| BMI | 30.3 ± 7.8 | 29.7 ± 7.9 | 31.4 ± 7.7 | 31.6 ± 7.6 | 31.2 ± 7.1 | <0.001 |
| Current smoking | 22.4 | 22.4 | 21.3 | 22.1 | 24.2 | <0.001 |
| Vitals at Index Visit |  |  |  |  |  |  |
| Pulse | 77.4 ± 11.6 | 77.3 ± 11.4 | 77.9 ± 11.5 | 77.7 ± 11.8 | 76.9 ± 12.2 | <0.001 |
| Systolic BP | 125 ± 16.1 | 120 ± 13.6 | 129 ± 14.7 | 133 ± 16.3 | 141 ± 18.7 | <0.001 |
| Hypertension (treated) | 22.5 | 11.6 | 31.5 | 44.9 | 63.4 | <0.001 |
| Current Medications (index visit) |  |  |  |  |  |  |
| Aspirin | 14.2 | 67.0 | 20.4 | 28.8 | 41.7 | <0.001 |
| Statin | 17.1 | 8.6 | 25.7 | 34.8 | 46.2 | <0.001 |
| Zetia | 0.9 | 0.9 | 1.0 | 0.9 | 0.8 | 0.69 |
| PCSK9 | 8 (0.01%) | 6 (0.01%) | 0 (0.00%) | 2 (0.01%) | 0 (0.00%) | 0.64 |
| Cholesterol Panel |  |  |  |  |  |  |
| Cholesterol | 196 ± 36.9 | 192 ± 34.6 | 208 ± 38.3 | 206 ± 39.8 | 200 ± 41.0 | <0.001 |
| HDL-c | 57.0 ± 15.6 | 58.7 ± 15.2 | 54.2 ± 15.4 | 53.2 ± 15.6 | 53.4 ± 15.9 | <0.001 |
| LDL-c | 114 ± 32.4 | 111 ± 30.3 | 125 ± 33.9 | 121 ± 35.4 | 114 ± 36.6 | <0.001 |
| LDL≥190 | 2.2 | 1.3 | 3.8 | 4.1 | 3.6 | <0.001 |
| Diabetes Mellitus | 9.6 | 3.4 | 10.1 | 20.4 | 43.8 | <0.001 |
| ELIX Comorbidity Score | 0.14 ± 3.7 | -0.14 ± 3.4 | 0.27 ± 3.9 | 0.69 ± 4.1 | 1.31 ± 4.3 | <0.001 |
| Department Specialty |  |  |  |  |  | <0.001 |
| Cardiology | 5.1 | 3.2 | 6.4 | 8.6 | 12.6 |  |
| PCP | 94.9 | 96.8 | 93.6 | 91.4 | 87.4 |  |

**Table IIIa. Baseline Characteristics in Women, by ASCVD Risk.** Variables expressed as frequency or mean ± standard deviation, where appropriate. Abbreviations: BP-blood pressure, ELIX- Elixhauser Comorbidity Index, PCP- primary care physician, PCSK-proprotein convertase subtilisin/kexin type 9 serine protease

| ASCVD Risk | Total  (123,198) | Low  (56,511) | Borderline  (13,981) | Intermediate  (36,602) | High  (16,104) | P-Value |
| --- | --- | --- | --- | --- | --- | --- |
| Age | 49.5 ± 13.7 | 38.4 ± 9.1 | 51.3 ± 6.9 | 58.1 ± 7.6 | 67.4 ± 7.7 | <0.001 |
| Age≥ 75 | 2.2 | 0.0 | 0.0 | 0.2 | 16.7 | <0.001 |
| Race |  |  |  |  |  | <0.001 |
| White | 88.3 | 86.9 | 89.1 | 89.5 | 89.9 |  |
| Black | 6.7 | 5.3 | 8.0 | 8.0 | 7.8 |  |
| Other | 5.0 | 7.8 | 3.0 | 2.6 | 2.3 |  |
| BMI | 30.4 ± 6.3 | 29.9 ± 6.6 | 30.7 ± 6.2 | 30.7 ± 6.1 | 31.1 ± 6.0 | <0.001 |
| Current smoking | 25.9 | 23.3 | 23.8 | 26.8 | 35.1 | <0.001 |
| Vitals at Index Visit |  |  |  |  |  |  |
| Pulse | 76.3 ± 11.7 | 76.3 ± 11.6 | 76.6 ± 11.4 | 76.5 ± 11.7 | 75.7 ± 11.9 | <0.001 |
| Systolic BP | 129 ± 15.6 | 124 ± 13.2 | 128 ± 14.4 | 132 ± 15.5 | 140 ± 17.8 | <0.001 |
| Hypertension (treated) | 27.8 | 12.2 | 25.7 | 37.8 | 61.1 | <0.001 |
| Current Medications  (index visit) |  |  |  |  |  |  |
| Aspirin | 18.0 | 5.6 | 16.8 | 26.8 | 42.7 | <0.001 |
| Statin | 19.5 | 7.8 | 18.9 | 27.8 | 42.3 | <0.001 |
| Zetia | 1.0 | 1.0 | 0.9 | 1.0 | 1.0 | 0.93 |
| PCSK9 | 3 (0.00) | 1 (0.00) | 1 (0.01) | 1 (0.00) | 0 (0.00) | 0.62 |
| Cholesterol panel |  |  |  |  |  |  |
| Cholesterol | 191 ± 36.1 | 188 ± 33.9 | 196 ± 35.9 | 196 ± 37.4 | 188 ± 38.8 | <0.001 |
| HDL-c | 46.6 ± 13.1 | 47.8 ± 13.0 | 47.1 ± 13.6 | 45.9 ± 13.0 | 43.2 ± 12.4 | <0.001 |
| LDL-c | 116 ± 32.5 | 113 ± 30.7 | 118.9 ± 31.6 | 119.2 ± 33.4 | 111 ± 35.6 | <0.001 |
| LDL≥190 | 2.0 | 1.3 | 2.3 | 2.8 | 2.4 | <0.001 |
| Diabetes Mellitus | 11.0 | 3.0 | 6.1 | 11.7 | 41.6 | <0.001 |
| ELIX Comorbidity Score | 0.45 ± 3.5 | 0.05 ± 3.0 | 0.32 ± 3.4 | 0.66 ± 3.7 | 1.46 ± 4.2 | <0.001 |
| Department Specialty |  |  |  |  |  | <0.001 |
| Cardiology | 5.4 | 2.9 | 5.0 | 7.0 | 11.2 |  |
| PCP | 95.0 | 97.1 | 95.0 | 93.1 | 88.8 |  |

**Supplemental Table IIIb. Baseline Characteristics in Men, By ASCVD Risk.** Variables expressed as frequency or mean ± standard deviation, where appropriate. Abbreviations: BP-blood pressure, ELIX- Elixhauser Comorbidity Index, PCP- primary care physician, PCSK-proprotein convertase subtilisin/kexin type 9 serine protease.
